# Geo-temporal distribution of 1,688 Chinese healthcare workers infected with COVID-19 in severe conditions – a secondary data analysis

**DOI:** 10.1101/2020.03.19.20032532

**Authors:** Wayne Gao, Mattia Sanna, Chi Pang Wen

**Author notes:** **Address correspondence to:** Wayne Gao, Master’s Program in Global Health and Development, Taipei Medical University - 250 Wuxing Street, 110 Taipei City, Taiwan. # 1323.

## Abstract

**Introduction:** The COVID-19 outbreak is posing an unprecedented challenge to healthcare workers. This study analyzes the geo-temporal effects on disease severity for the 1,688 Chinese healthcare workers infected with COVID-19.

**Method:** Using the descriptive results recently reported by the Chinese CDC, we compare the percentage of infected healthcare workers in severe conditions over time and across three areas in China, and the fatality rate of infected healthcare workers with all the infected individuals in China aged 22-59 years.

**Results:** Among the infected Chinese healthcare workers whose symptoms onset appeared during the same ten-day period, the percentage of those in severe conditions decreased statistical significantly from 19.7% (Jan 11 – 20) to 14.4% (Jan 21 – 31) to 8.7% (Feb 1 – 11). Across the country, there was also a significant difference in the disease severity among patients symptoms onset during the same period, with Wuhan being the most severe (17%), followed by Hubei Province (10.4%), and the rest of China (7.0%). The case fatality rate for the 1,688 infected Chinese healthcare workers was significantly lower than that for the 29,798 infected patients aged 20-59 years—0.3% (5/1,688) vs. 0.65% (193/29,798), respectively.

**Conclusion:** The disease severity improved considerably over a short period of time in China. The more severe conditions in Wuhan compared to the rest of the country may be attributable to the draconian lockdown. The clinical outcomes of infected Chinese healthcare workers may represent a more accurate estimation of the severity of COVID-19 for those who have access to quality healthcare.

## Introduction

The first cluster of novel coronavirus disease 2019 (COVID-19) originated in Wuhan, China on December 31, 2019^1^. By February 27, 2020, 82,170 cases of COVID-19, including 2,804 deaths, had been confirmed, mainly in China and more than 40 countries and areas^2^. Severity and transmissibility of the COVID-19 have been the most important issues researchers and public health authorities have been urgently trying to tackle since the start of the outbreak. The first studies reporting single-center case series of hospitalized patients of COVID-19 showed high rates of admission to intensive care (32%), high mortality (15%)^3^, and case fatality rates (CFR) of 4.3% (6/138)^4^ and 11% (11/99)^5^ in Wuhan, while lower figures were reported outside the city^6^. However, the high CFRs observed in Wuhan are likely to be overestimated, as the cited studies have mainly considered patients with severe symptoms who were hospitalized, excluding mild and asymptomatic patients who are less likely to be admitted to the hospital^7^. The most recent estimation of an overall 2.3% CFR reported by China CDC^8,9^ is thus tentative, and more specific figures will remain undetermined until a later point.

Frontline healthcare workers are significantly more likely than general public to come into contact with the infected individuals. Knowing the clinical outcomes of healthcare workers who have been infected may provide critical information on both risk and disease severity, especially when proper healthcare services are available.

## Methods

We use the descriptive results from “The epidemiological characteristics of an outbreak of 2019 novel coronavirus disease”, published by the Chinese CDC^8^, which is, by far, the most comprehensive epidemiological investigation of COVID-19 in China. This report covers all the 72,314 COVID-19 cases and the 1,688 laboratory-confirmed cases among healthcare workers as of February 11. A similar version in English was also published in the Journal of American Medical Association(JAMA)^9^, including clinical outcomes, classified by severity (mild symptoms vs. severe/critical conditions) and by time into three 10-day periods based on the onset of symptoms reported by the patients. Using chi-squared test and Fisher’s exact test, we analyze the changes in percentage of infected healthcare workers in severe conditions over the same three periods (1/11-1/20, 1/21-1/31, and 2/1-2/11) and across the same three regions (Wuhan City, Hubei Province excluding Wuhan, China excluding Hubei). We also compare the CFR for all infected patients aged 20-59 and for healthcare workers.

## Results

The CFR for the 1,688 infected Chinese medical workers was significantly lower than the CFR for the 29,798 infected patients aged 20-59 years—0.3% (5/1,688) vs. 0.65% (193/29,798), respectively. As shown in Table 1, for infected healthcare workers, the percentage of those in severe conditions decreased significantly, from 19.7% (Jan 11 – 20) to 14.4% (Jan 21 – 31) to 8.7% (Feb 1 – 11).

**Table 1.** Statistical analysis of COVID-19 cases among healthcare workers in China.

| Dates | Wuhan |  |  | Hubei except Wuhan |  |  | China except Hubei |  |  | Fisher's exact test |
| --- | --- | --- | --- | --- | --- | --- | --- | --- | --- | --- |
|  | Confirmed cases | Severe cases | (%) | Confirmed cases | Severe cases | (%) | Confirmed cases | Severe cases | (%) |  |
| **Jan 11-20** | 233 | 52 | (22.3) | 48 | 8 | (16.7) | 29 | 1 | (3.5) | 0.001 |
| **Jan 21-31** | 656 | 110 | (16.8) | 250 | 29 | (11.6) | 130 | 10 | (7.7) | 0.01 |
| **Feb 1-11** | 173 | 22 | (12.7) | 95 | 3 | (3.2) | 54 | 3 | (5.6) | 0.001 |
| **Chi-square test** |  |  | 0.03 |  |  | 0.0004 |  |  | 0.07 |  |
| **Total** | 1062 | 184 | (17.3) | 393 | 40 | (10.2) | 213 | 14 | (6.6) | <0.0001 |

Considering only patients whose symptom onset appeared during the same 10-day period, the percentage of those in severe conditions exhibits a distinctive geographic distribution centered around the lockdown epicenter of the outbreak - Wuhan being the most severe (17%), followed by Hubei Province excluding Wuhan (10.4%), and by of China excluding Hubei (7.0%) (Figure 1). Of the 149 Chinese health workers who were infected with COVID-19 outside of Wuhan City after February 1, only 4% were in severe condition (6/149) and there were no deaths.

**Figure 1.**
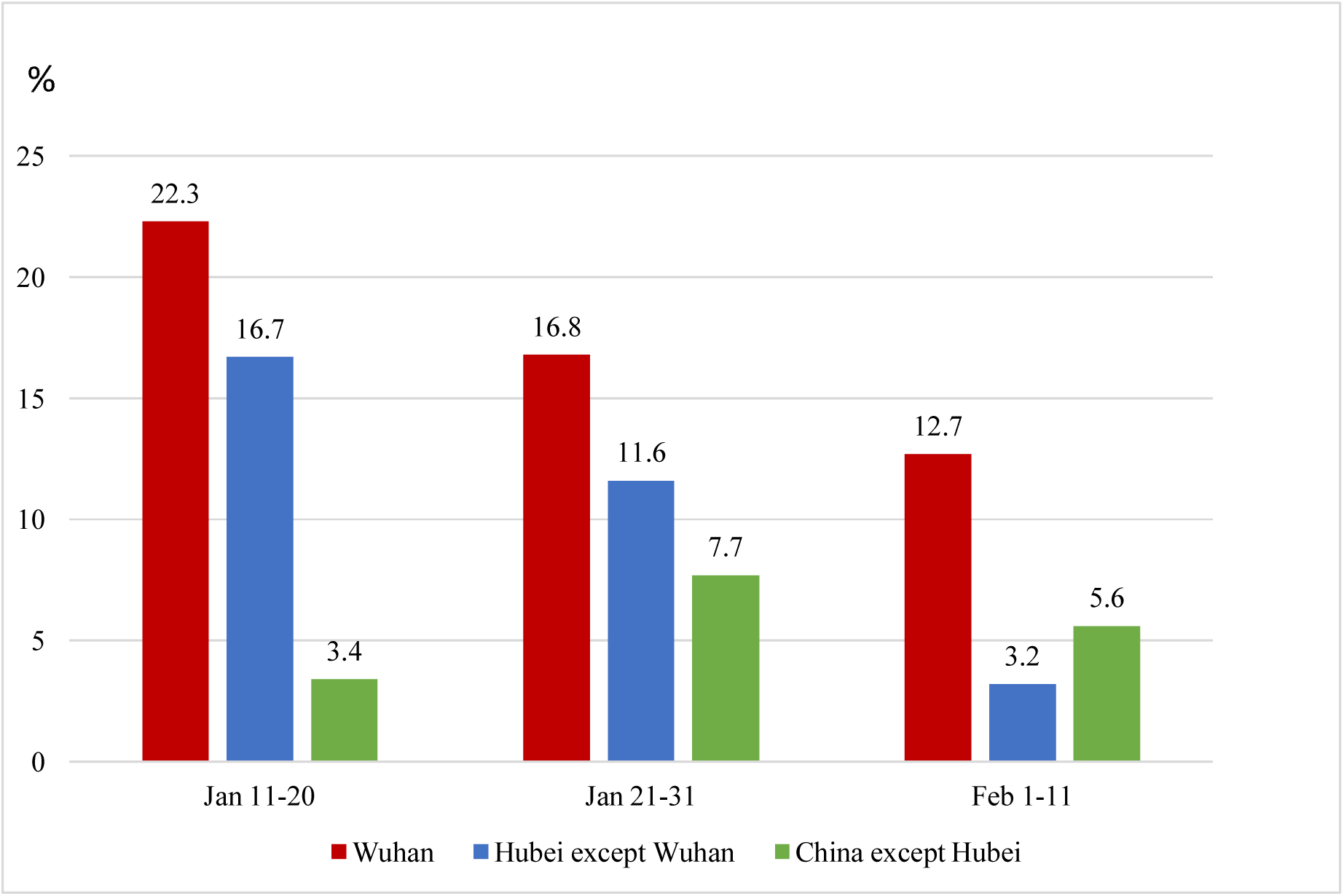
Percentage of healthcare infected workers in severe conditions by dates of symptoms onset regions in China.

## Discussion

Preventing nosocomial transmission is a top priority during an epidemic. The consistent higher percentage of COVID-19 infected healthcare workers in severe conditions in Wuhan may be related to the draconian city lockdown and the established of massive quarantines camps, since viral factors such as mutation or adaptation are unlikely to be responsible for the difference observed for the three 10-day periods (as they occur over longer periods) and across the three locations taken into account. The transmission control was improper and co-infections were common during the initial period of the outbreak in Wuhan. One recent study found a correlation between the higher CFR in Wuhan with the higher healthcare burden compared with other provinces in China^10^. It is unthinkable that sickening citizens, infected or not, and family members were confined in a city lockdown with an overwhelmed medical system and that they were forbidden from going to other provinces to seek for better care to save their lives. This also inevitably created logistical as well as psychological impact on healthcare workers operating in those harsh environments, described in a call for international assistance by Chinese medical workers, as “nurses’ mouths are covered in blisters and some nurses have fainted due to hypoglycaemia and hypoxia^11^. Sixty-three percent of the infected healthcare workers have been in Wuhan, including the 5 who died as a result of the infection^9^. However, the 0.3% CFR for the 1,688 medical workers (who were mainly aged less than 60 years as indicated in the report^8^), may represent a proper estimation of the severity of COVID-19 in this age group in countries with universal access to quality medical services. Our study has a few limitations: There may be infected healthcare workers who still have not completed their natural progression of the disease with definite outcomes. However, as of February 28, no additional deaths of healthcare workers were reported. Second, our data analysis is based on a governmental investigation of the outbreak, thus the validity of the descriptive results in the report inevitably determines the accuracy of our secondary analysis.

The spectrum of clinical severity of a novel communicable disease is critical in knowing the potential impact of an ongoing epidemic. Our finding that the spectrum of COVID-19 severity decreases eccentrically from the epicenter as well as over short periods of time, will help to avoid drastic, costly, and fear-driven measures that impede not only daily activities, but also a proper containment of the epidemic.

## Data Availability

The data are all available

## Notes

The authors declare that they have no conflicts of interest and funding.

### Competing Interest Statement

The authors have declared no competing interest.

### Funding Statement

No funding

